# Design and Evaluation of a Novel Thrombectomy Device

**DOI:** 10.64898/2026.01.05.26343477

**Authors:** Łucja Aleksandra Żurawska, Mario van der Wel, David Jager, Remi van Starkenburg, Paul Breedveld, Frank Gijsen

## Abstract

Deep vein thrombosis is a disease that occurs when a blood clot is formed in a vein and occludes the vessel lumen, blocking the blood flow, causing pain and even disability and possibly leading to complications such as postthrombotic syndrome and pulmonary embolism. Treatments for DVT include mechanical thrombectomy: introducing a device into the vasculature to remove thrombus. Currently used devices either macerate the thrombus to aid removal or pierce the thrombus to reach its distal side. This can pose risk of fragmentation or distal embolization, or in case of fibrous, cohesive thrombi can be hard to achieve due to their resistance to deformation. The following study proposes an alternative approach of bypasing the thrombus via the space between the thrombus and the vessel wall in order to avoid thrombus penetration. The design implements a strategy of simultaneously gripping the clot and expanding the vessel lumen in order to create space between the thrombus and the vessel wall while advancing along the clot’s length in incremental steps. The prototype has been evaluated in a custom-made experimental setup using phantom vessels and thrombi analogs. The proof-of-concept experiments have shown that the device can successfully bypass and in some cases even remove thrombi. The study shows promising results for this new kind of device and can be a foundation for future research into applying similar removal strategies in thrombectomy.

## Introduction

Deep vein thrombosis (DVT) occurs when a blood clot forms in the vasculature, obstructing the vein’s lumen and preventing blood flow. It can lead to postthrombotic syndrome [1], causing long term disability and in some cases the clot can dislodge and travel to the lung vasculature, causing pulmonary embolism [2]. Currently treatments include compression stockings, anticoagulation, thrombolysis and mechanical thrombectomy [3]. During a mechanical thrombectomy a device capable of capturing the thrombus is inserted into the vasculature using a catheter. During the procedure it is important to not damage the vessel wall, which can lead to further thrombogenesis or haemorrhage and to avoid thrombus fragmentation, which can lead to longer intervention time and distal embolization. Mechanical thrombectomy in case of DVT can include first breaking up the clot and then removing it using negative pressure such as in case of rheolytic thrombectomy (e.g. by using the AngioJet System, commercialised by Boston Scientific) where a stream of saline is used for maceration and aspiration [4]. Another available technique would be using a device such as the ClotTriever System (Inari Medical) [5], Fig. 1. The device constitutes of a mesh collection bag which can be collapsed to very small diameter. The mesh is collapsed within a microcatheter when introduced into the vasculature. The microcatheter penetrates the thrombus in order to reach its distal end. Once it is placed there, the microcatheter is pulled back, unsheathing the mesh collection bag which expands to the diameter of the vessel lumen. The bag is then pulled over the thrombus, allowing its removal.

**Fig 1.**
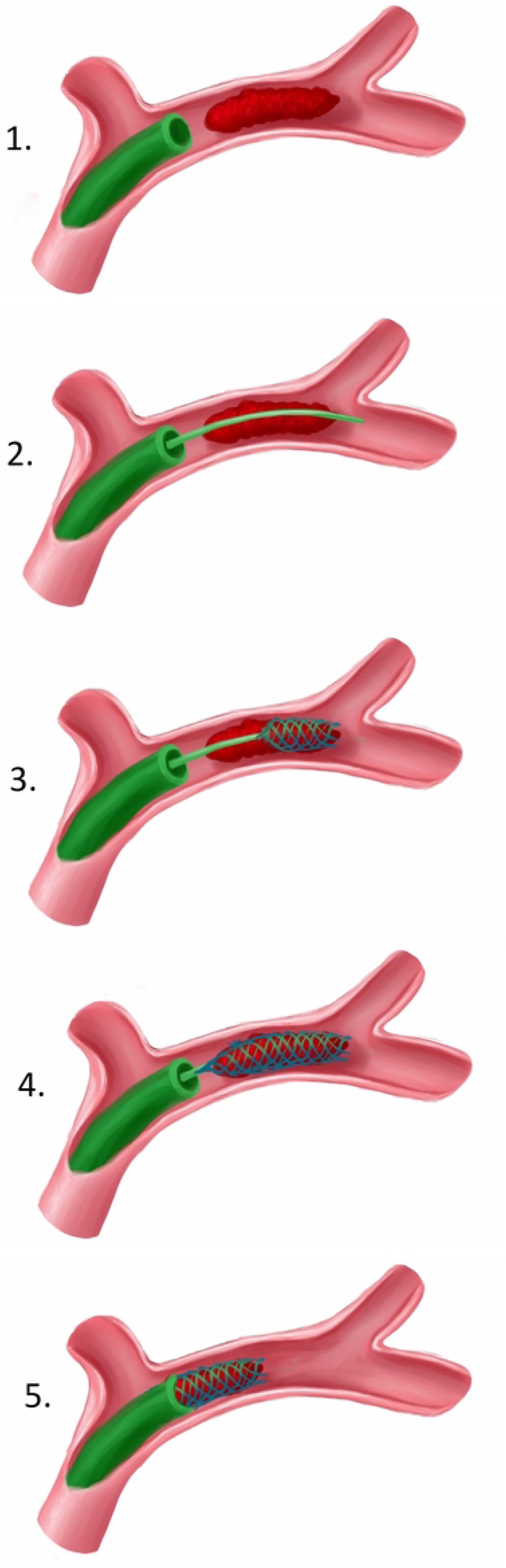
Example of a currently used thrombectomy device. 1: A catheter is placed at the site of occlusion. 2: A microcatheter, containing a collapsed mesh bag penetrates the thrombus. 3: The microcatheter is pulled back, unsheathing the mesh bag. 4: The mesh bag radially expands, capturing the thrombus when pulled over it. 5. The captured thrombus is removed.

While these methods are quite effective they still pose some risks, which are dependent on the type of thrombus they are used for. Thrombi can have various mechanical properties depending on their composition [6]. Thrombi primarily consist of platelets, red blood cells (RBC) and fibrin. Thrombi with a high concentration of red blood cells are rather soft and easily broken. Thrombi with a high platelet concentration are firm and less fragile than the RBC-rich ones. Platelet-rich thrombi are also often interwoven with fibrin which makes them firmer. Fibrin can also bond to the vessel walls, which is important in the context of removal [7]. RBC-rich thrombi are easily fragmented and even the insertion of a catheter can cause it to fragment, which can lead to distal embolization. Fibrous, platelet-rich thrombi on the other hand are harder to penetrate or break down and their bond with the vessel wall can resist removal and cause strain on the vessel wall during a removal attempt.

The goal of this research is to create a method of mechanical thrombectomy that does not require the maceration or penetration of the thrombus. This study presents a proof-of-concept design of a device designed to reach the distal end of the thrombus via the space between the thrombus and the vessel wall. A prototype was made based on the design and evaluated in an experiment.

## Design

### Concept design

In order to reach behind the thrombus without penetrating it the device has to be inserted between the thrombus and the vessel wall. The thrombus is usually in contact with the vessel wall so inserting a tool between them could scrape the thrombus and cause fragmentation or damage the vessel lining. To prevent that the vessel lumen can be slightly expanded, thus breaking the contact between the thrombus and the vessel wall. This can be achieved by exerting an outward radial force on the inner circumference of the vessel wall. However exerting an outward force along the entire length of the thrombus could potentially lead to the thrombus dislodging and getting pushed away by the force of the blood flow. In order to prevent this the device can comprise of two parts: while one part expands the vessel lumen, the other one maintains the grip on the thrombus to prevent its dislodgement. In the procedure to capture the thrombus entirely, the first part of the device first advances along an increment of the thrombus length while expanding a small portion of the vessel and then grips that small portion of the thrombus thus “replacing” the force of the vessel wall that was holding the thrombus in place. Next the second part of the device expands and advances an increment further and grips the thrombus there. Once the second part establishes the grip, the first part can release the grip, expand the vessel and advance further. This incremental action is repeated until the end of the thrombus is reached. The working principle is illustrated in Fig. 2. The two parts can be grippers comprising of a set of fingers. The fingers moving radially inward exert force on the thrombus and grasp it. The fingers moving radially outward exert force on the vessel and expand it. While the minimal amount of fingers needed to grasp the thrombus or expand the vessel is two, three fingers per part have been chosen for this design to ensure a more stable grip and more uniform expansion of the vessel lumen. In order to allow the advancement of the expanding set of fingers past the grasping set of fingers, the fingers are arranged as shown in Part 5 of Fig. 2.

**Fig 2.**
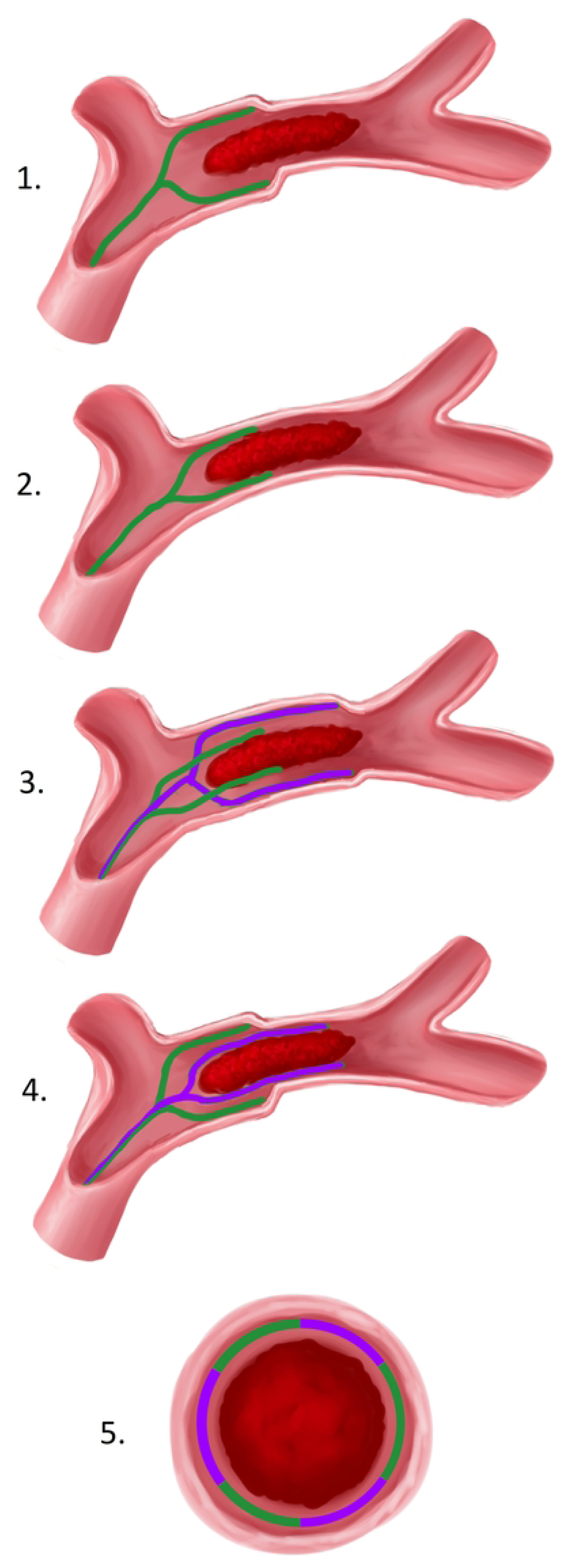
Working principle of the device and the placement of the grippers’ fingers. 1: The green part expands the first increment of the vessel lumen. 2: The green part closes in and grasps the thrombus. 3: The purple part expands the lumen an increment further than the green part. 4: The purple part closes in and grasps the thrombus, the green part releases its grip and can now advance and expand the vessel lumen an increment further than the purple part. The cycle is repeated until the end of either part reaches the end of the thrombus. 5: Cross section view of the vessel. Green: one set of fingers, purple: second set of fingers.

### Final design

The diameter of the device has been set to 1 cm as that size is comparable to the vessel lumen diameter of leg veins [8]. The length of the device’s fingers has been set to 4 cm. This length has been chosen so that a 2 cm long thrombus can be removed even if some displacement occurs. Because our design was intended to evaluate our incremental expand, grip and release method to remove thrombi, we decided to design only the end-effector of an envisioned surgical device, meaning that only the fingers and their actuation system were developed, not a connection with a semi-flexible catheter-like shaft that is inserted through the skin. However, the fingers were designed in such a way that they are long and very slender and therefore suited for future compression like a stent and actuation from the proximal handle side of the semi-flexible shaft, when they serve as end-effector of a surgical device that is inserted through the patient’s skin.

Each of the fingers of the device has to be able to move inwards and outwards. In order to avoid the complexity of hinged joints and their actuation a compliant mechanism has been chosen. Each finger was made of 0.3 mm diameter U-shaped spring steel wire bent to a slight S-shape in side view (Fig. 3). The two ends of each wire are placed within two stiff, steel tubes of 0.4 mm inner diameter and 0.9 outer diameter. It has been empirically found that an approximate 30 degrees angle of the bend in the S-shape should in principle be enough to provide sufficient force to expand the vessel wall while at the same time still allowing the tubes to slide over the bend with little friction. When the tubes are covering only the straight ends of the wire the finger is in its default, expanding position, top part of Fig. 3. In order to switch to the grasping position the tubes are slid over the proximal bends of the wire, which act as pre-bent flexures. The tubes deform the flexures and the end of the finger switches to the opposite, gripping position, shown in bottom part of Fig. 3.

**Fig 3.**
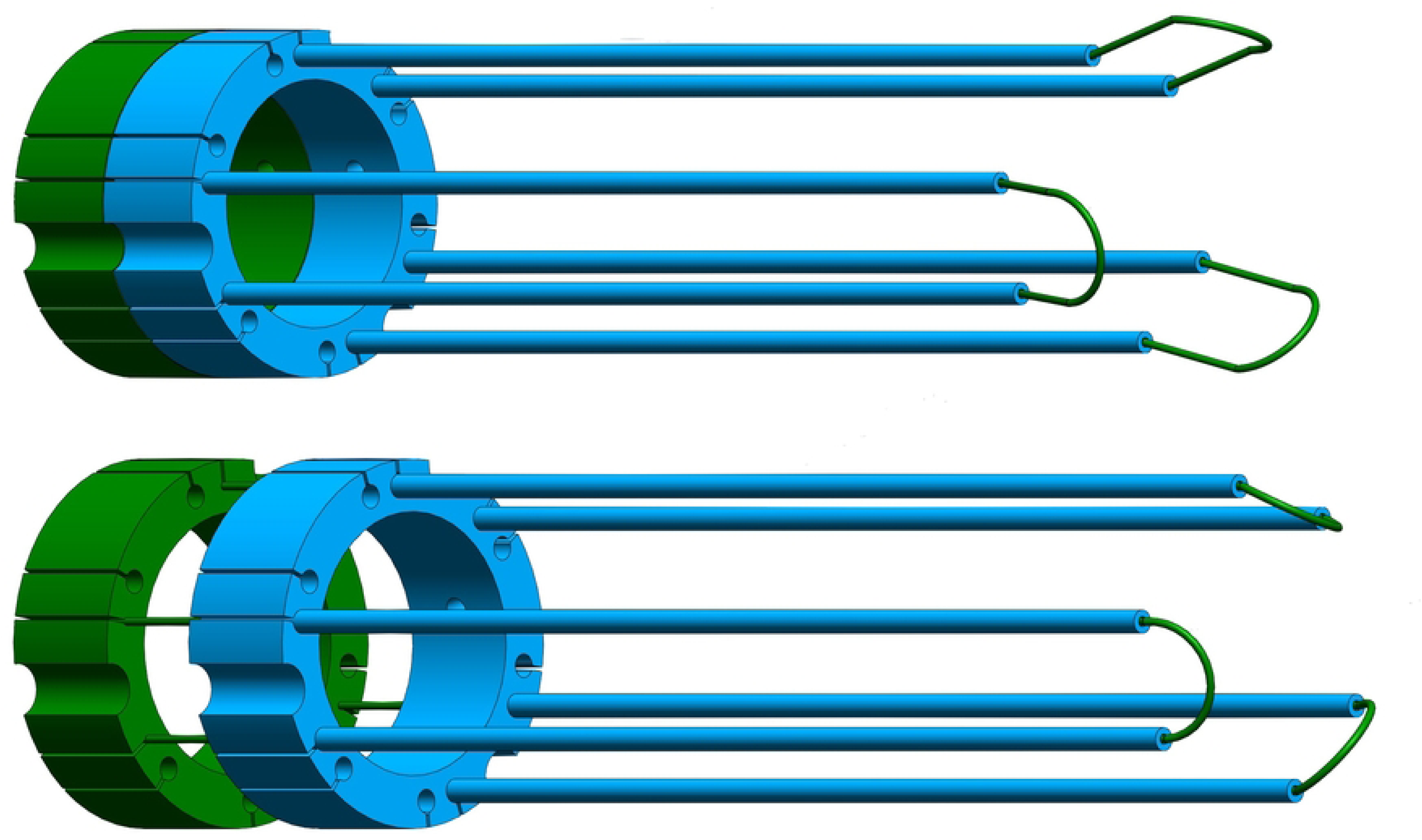
One set of fingers of the device. The three U-shaped spring steel wires (green) are connected by a ring (green) and the six rigid tubes (blue) are connected by another ring (blue). Top: the rings are at a close distance from each other, the proximal halves of the S-shaped curves of the wires are released and the fingers are in the expanding position. Bottom: the rings are at a larger distance from each other, the proximal curves of the wires are constrained and the wires are deformed to the gripping position.

With this design the device contains two sets of three wires actuated by two sets of six tubes. Since all the elements per set perform the same movements throughout the device’s operation they are rigidly connected by one ring per set, Fig 3. In order to allow the sets of fingers to pass by each other i.e. allow the expanding set to advance from behind the grasping set to the front, each set occupies half of the circumference of the device. The rings connecting one set of wires or tubes have slots, visible in Fig 3, through which the tubes of the second set can slide.

All the tubes and wires of the device need to move in a specific manner relative to each other to achieve the desired operation of the device. The movement that each set of wires and tubes performs throughout the cycle is presented in Table 1.

**Table 1.** Movement of each group of elements throughout one cycle of the device’s operation, see also Fig. 3-4.

|  | purple wires | orange tubes | green wires | blue tubes |
| --- | --- | --- | --- | --- |
| Set 1 (blue and green): transition to expanding<br>Set 2 (orange and purple): transition to gripping | static | advancing | static | retracting |
| Set 1: expanding and advancing<br>Set 2: gripping and static | static | static | advancing | advancing |
| Set 1: transition to gripping<br>Set 2: transition to expanding | static | retracting | static | advancing |
| Set 1: gripping and static<br>Set 2: expanding and advancing | advancing | advancing | static | static |

To achieve this desired movement the four rings connecting the wires and tubes need to be actuated in a specific sequence of axial translations. To avoid rotation of the rings during their forward and backward motion, the rings are constrained by a frame, grey in Fig. 4. The rings have semicircular grooves in their outer circumference. In these grooves rods are placed that are connected to the outer frame of the device, allowing forward and backward translation of the rings while restricting rotation of the rings. The rings are actuated by a cam, yellow in Fig. 4, which is placed inside the rings. The cam is a rotating cylinder with four grooves. The grooves are graphical representations of one movement cycle of each ring, as presented in Table 1, and wrapped around the circumference of the cylinder (Fig. 4). Each of the four rings is equipped with a pin protruding inward and sliding through one of the cylinder’s grooves as it is rotated, producing a back and forth axial ring motion.

**Fig 4.**
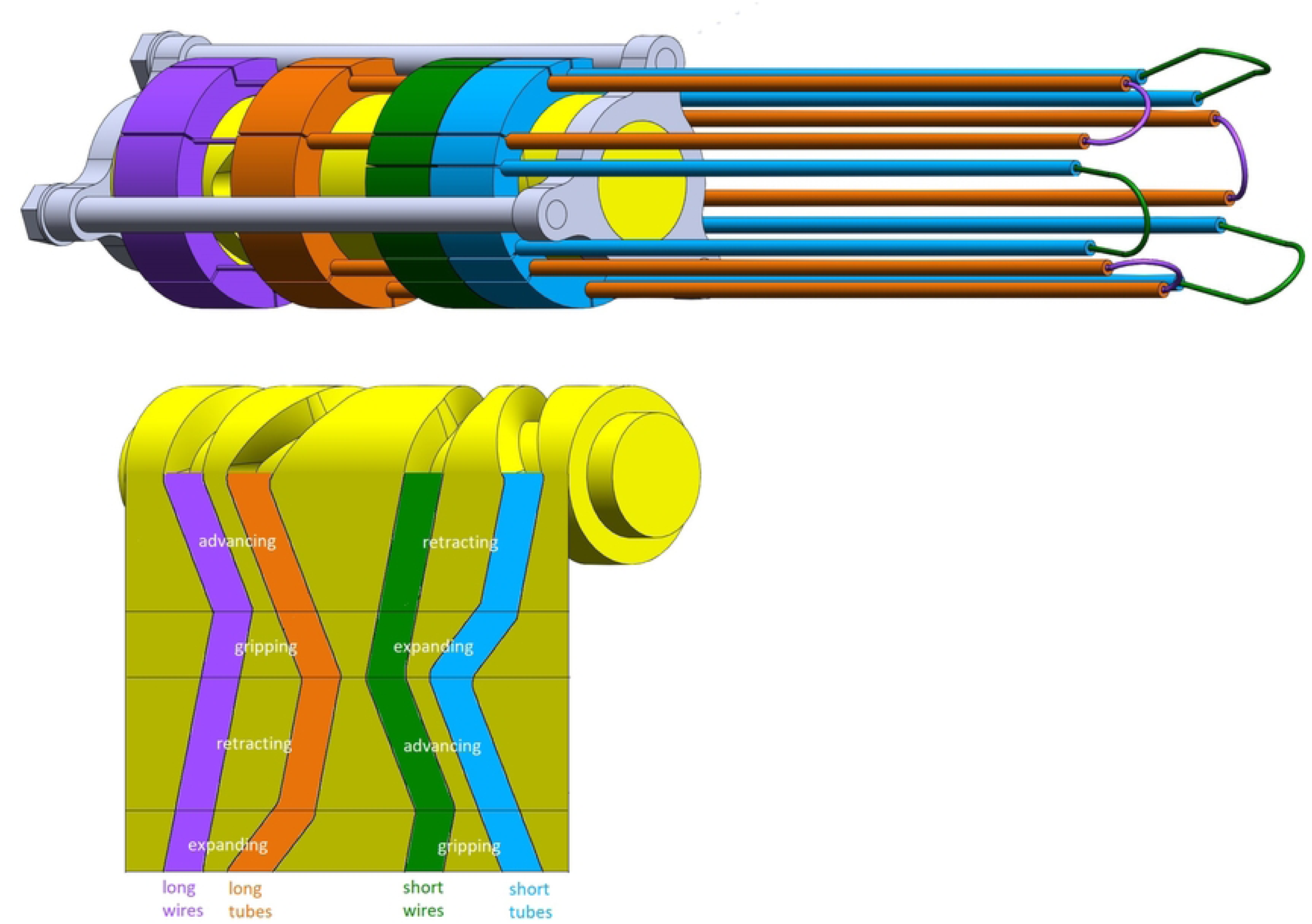
Outer frame with both sets of arms and grooved cam cylinder. Top: grey frame placed around the rings and yellow cam-cylinder with four cam paths placed inside the rings. Bottom: unwrapped view of the cam paths in the cylinder with indication of phase of movement of each set of arms.

The grooves on the cam cylinder allow back and forth axial motion of the rings, but after each rotation of the cylinder the rings return to their initial position. For a correct operation of the device the fingers have to advance when expanding the vessel lumen and stay static when grasping the thrombus. In order to allow this net forward motion the cam is additionally equipped with a worm screw. The rotation of the screw simultaneously actuates the cam-cylinder and pushes it forward so that the fingers advance over the thrombus at the correct speed, Fig. 5. The frame of the device (grey) is placed on a slider rail (white) allowing only axial translation. The cam cylinder is rigidly connected to the worm screw which rotates through a nut that is located at one end of the slider rail. The threading of the nut was chosen such that it provides an advancement of 2 mm per rotation, allowing the desired movement of the fingers. The worm screw is driven by a manually actuated knob (yellow). Fig. 5 shows the final design in assembled and exploded view.

**Fig 5.**
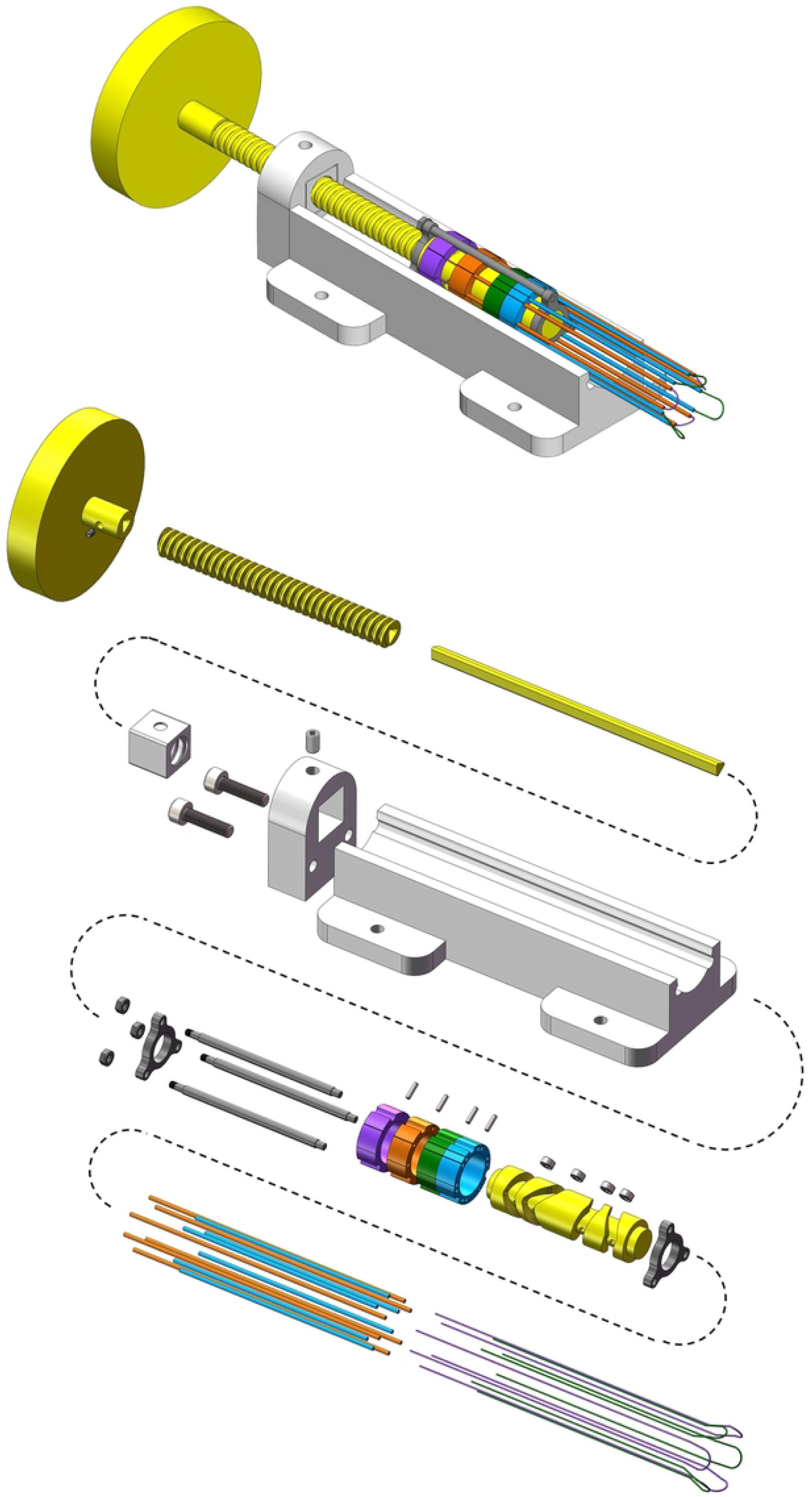
Complete device. Top: assembled view. Bottom: exploded view. First row, from left to right: knob, worm screw steel core, worm screw thread (yellow). Second row: threaded nut insert, nut enclosure, slider rail (white). Third row: base of the frame, frame rods, rings and pins, cylinder cam and ball bearings, top of the frame. Fourth row: tubes, wires.

The prototype of the device was manufactured at the DEMO Central Workshop of TU Delft. The rings are made from bronze, generating low friction with the aluminum cylinder and frame and stainless steel tubes, allowing a smooth motion of the device. The pins of the rings were additionally equipped with 3 mm diameter ball bearings to provide near frictionless sliding along the cam cylinder’s grooves. The cam cylinder was created with a milling machine while the rings were made with electrical discharge machining. The worm screw was manufactured from an inner steel rod and outer 3D-printed thread. Similarly the nut comprises of a steel enclosure and 3D-printed threaded insert. Fig 6 shows the finished prototype. A video of the prototype being actuated can be found in supplemental material S1 Video.

**Fig 6.**
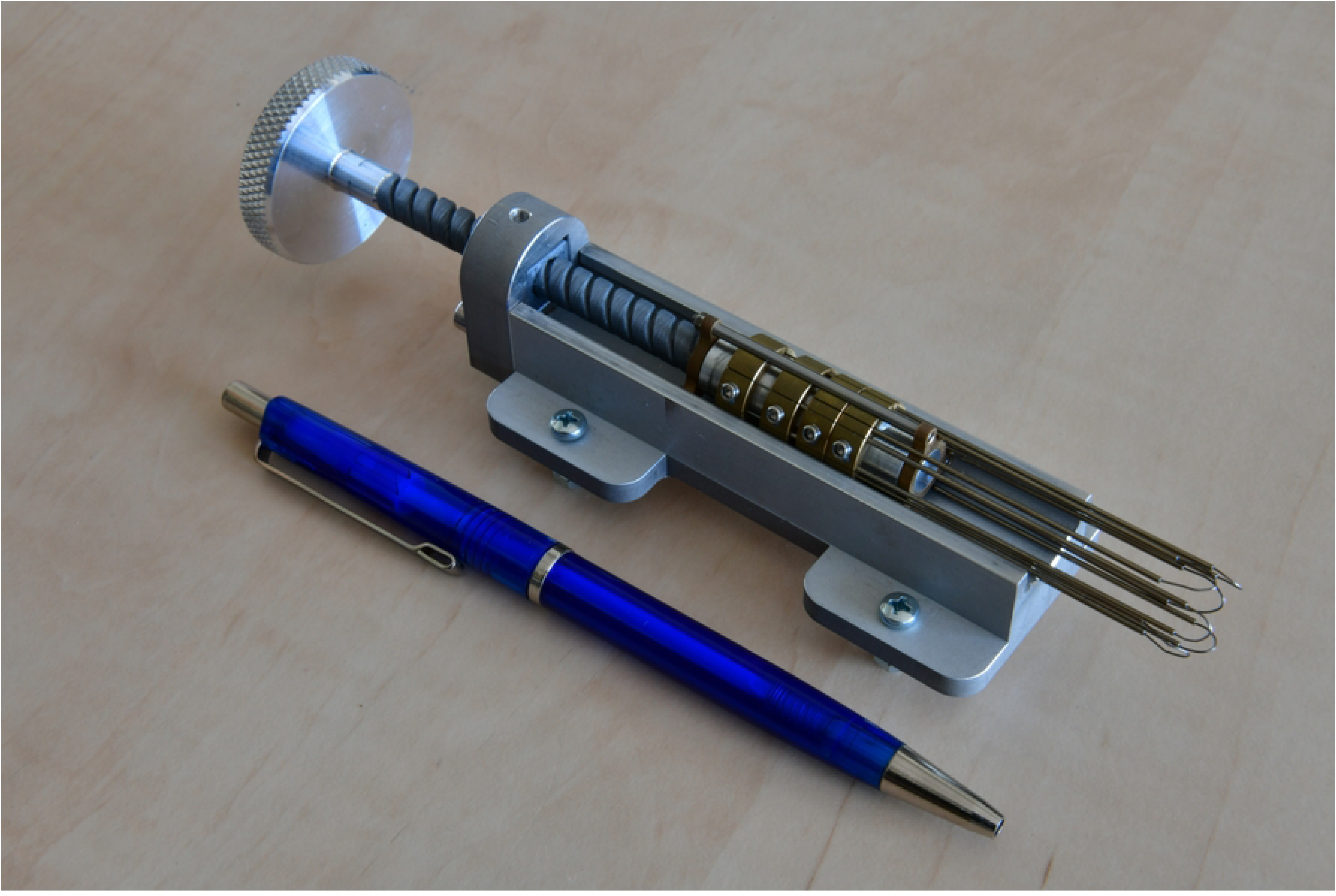
Photo of the finished prototype. The figure shows a pen to indicate the scale of the prototype.

## Evaluation

### Goal of the experiment

In a proof-of-principle experiment, the functioning of the developed prototype was evaluated in a custom-made experimental setup. The goal of the experiment was to evaluate whether the prototype fulfills the following functions:

- Gripping - does the thrombus remain gripped throughout the device’s operation? In the presence of a perfect grip, the thrombus would not slip while being gripped, and therefore not slide during the forward motion of the fingers. In case of a non-perfect grip there is a possibility that the thrombus will be pushed forward due to the friction between the thrombus and the advancing set of fingers. For that reason the distance over which the thrombus is pushed forward during the device’s operation can be treated as a measure of the efficiency of the grip.
- No fragmentation - after the removal of the device, are there any thrombus fragments remaining in the experimental setup? If so, what size?

### Experimental setup

The experimental setup, Fig. 7, consists of a test rig with a blood vessel phantom and a thrombus analog. The human tissue used in the research was obtained on 23 October 2024 and processed using a protocol approved by the Medical Ethical Committee at Erasmus MC (NL80622.078.22). Written consent was obtained from the tissue donor. The two types of thrombus analogs used in the experiment represent the two ends of the spectrum of thrombi’s mechanical properties: the fibrous, platelet-rich thrombi (Fig 7 top right) being the most cohesive and frictious type and the RBC-rich thrombi (Fig 7 middle right) representing the softer end of the properties spectrum. The thrombus analogs are cylindrical and have approximate diameter of 12 mm and approximate length of 24 mm. The thrombus analogs were manufactured from human blood according to the protocol used by Cahalane et al [9]. The blood vessel phantom in Fig. 7 is a tube manufactured from silicone. This material is often used to mimic the properties of blood vessels as it has similar elastic properties and its transparency allows observation of the prototype’s operation [10]. In order to test the device across a range of vessel properties two blood vessel phantoms were manufactured for the purpose of the experiment: the softer one made from A10 Shore hardness silicone and the stiffer one from A20 Shore hardness silicone. Each tube contains a flange at one end used for mounting within the experimental setup. The test rig was made from laser-cut PMMA sheets. The container consists of a reservoir and a slot. The reservoir is filled with HEPES buffer so that the experiments can be performed in wet conditions. The reservoir is equipped with a ruler so that the position of the thrombus analog can be recorded throughout the experiment. The slot is used to insert a PMMA sheet fastened to the prototype. A PMMA sheet with a circular hole is placed on top of the reservoir in order to place the phantom vessels within the reservoir.

**Fig 7.**
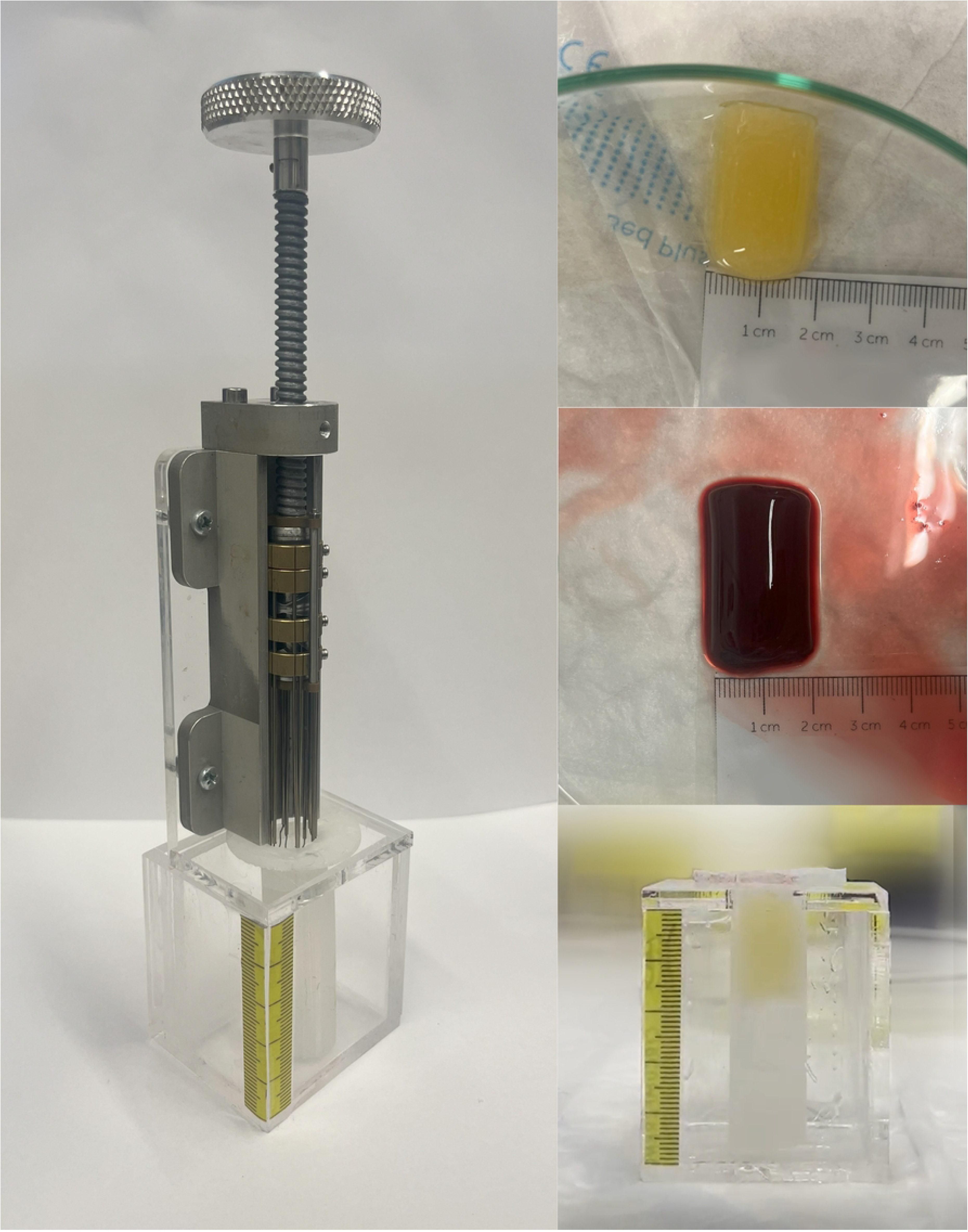
Experimental setup. Left: phantom vessel and prototype placed in the test rig. Top right: platelet-rich thrombus analog. Middle right: RBC-rich thrombus analog. Bottom right: platelet-rich thrombus analog placed in the phantom vessel placed in the test rig.

### Experimental protocol

For the purpose of the experiments two RBC-rich thrombus analogs and two platelet-rich thrombus analogs were produced. One RBC-rich thrombus analog was used in the experiment using the A10 hardness phantom vessel and the other RBC-rich thrombus analog was used in the experiment using the A20 hardness phantom vessel. The platelet-rich thrombus analogs were used likewise. In total four experiments were performed, each following the same procedure. A thrombus analog was placed inside a phantom vessel (as shown in Fig 7 bottom right), the arms of the device were placed at the opening of the phantom vessel (as shown in Fig 7 left), the device was manually actuated until its arms reached behind the thrombus analog (as shown in Fig 8 left) and lastly the device was removed from the phantom vessel. The position of the thrombus analogs before and after the the arms reaching behind was measured using the ruler placed on the reservoir in order to evaluate the efficiency of the grip. After removing the device the reservoir and phantom vessel were visually inspected for remaining thrombus analog fragments.

**Fig 8.**
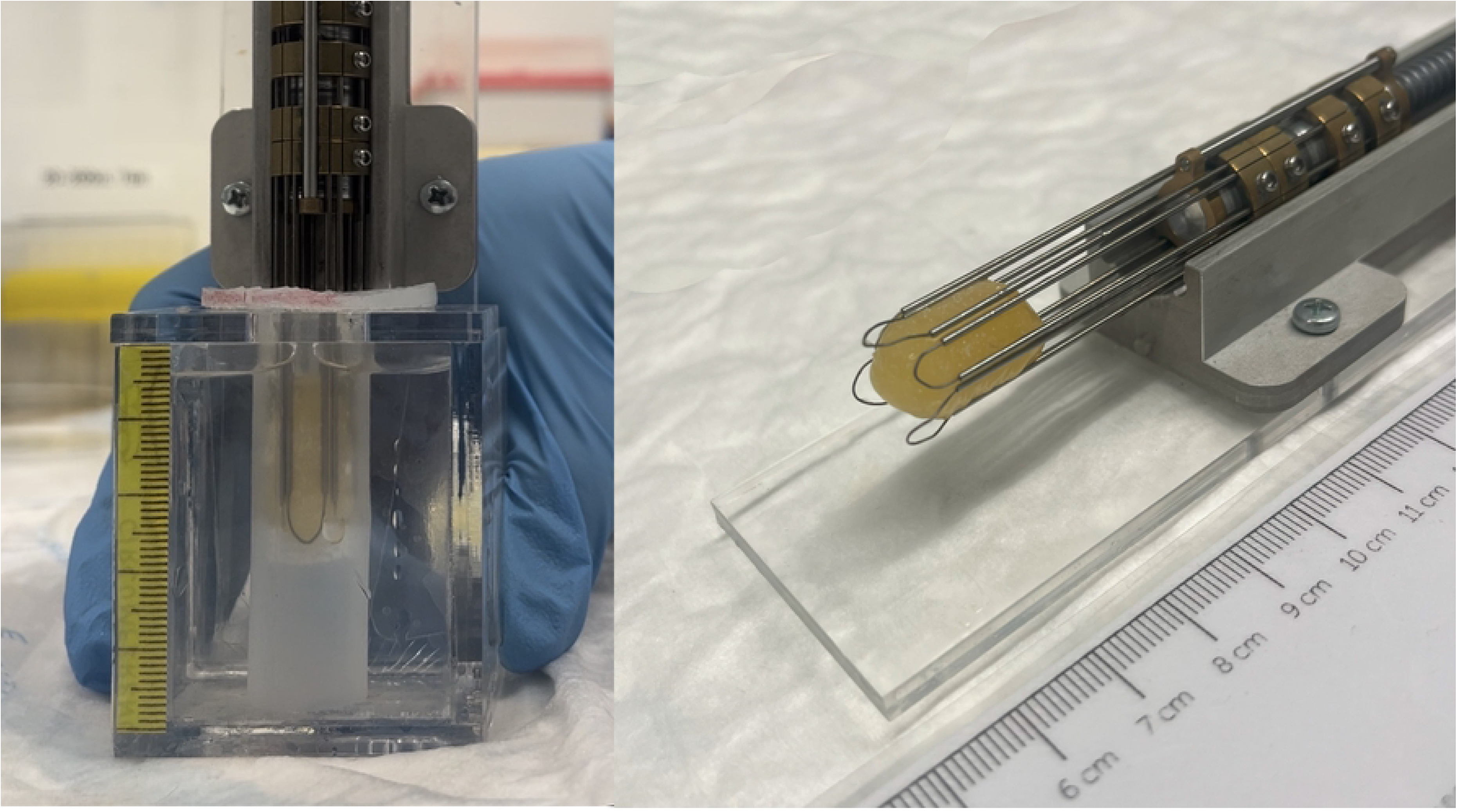
Photographs from the experiment using a platelet-rich thrombus analog. Left: grippers of the prototype reaching the end of the thrombus analog: moment after which removal begins. Right: platelet-rich thrombus analog gripped by the prototype after removal.

### Experimental results

The results of the experiments are presented in Table 2. In all of the experiments the end of the device reached the distal end of the thrombus analog. In the two experiments conducted on the platelet-rich thrombus analogs (in both phantom vessels) the prototype managed to successfully remove the thrombus analogs. It was observed, however, that the thrombus analogs were pushed downward during the advancement of the device’s arms. No signs of thrombus analog fragmentation were found in the test setup after the removal.

**Table 2.**
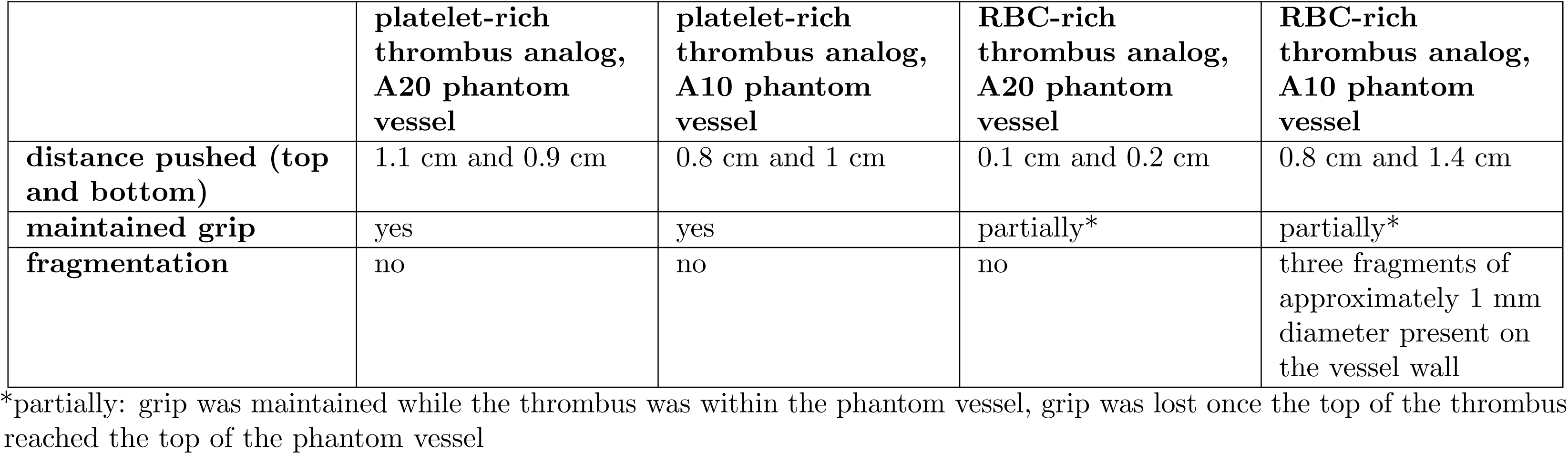
Results of the experiments.

In the two experiments conducted on the RBC-rich thrombus analogs the device did not manage to remove the thrombus analogs out of the phantom vessels, but it did maintain the grip over some distance during the removal. In both experiments the grip on the RBC-rich analogs was lost when the top of the thrombus analogs reached the opening of the phantom vessel. The thrombus analogs were pushed downward during the advancement of the device’s arms. When using the firmer phantom vessel no signs of fragmentation were observed within the test setup. When using the softer phantom vessel three small fragments of the RBC-rich thrombus analog (about 1 mm in diameter each) were found on the phantom vessel’s inner wall.

## Discussion

Currently used thrombectomy devices either break up the thrombus or penetrate it in order to reach behind it. This poses a risk of distal embolization which can lead to pulmonary embolism and increase the risk of post-thrombotic syndrome. The goal of this study was to develop a device that can bypass the thrombus without penetrating it in order to minimize the risk of fragmentation. The resulting design moves along the circumference of a thrombus incrementally while simultaneously exerting an outward radial force on the vessel wall in order to disturb the connection between the thrombus and the vessel wall and maintaining grip with the thrombus to avoid dislodgement. The prototype was tested in a custom-made setup using thrombus analogs and silicone phantom vessels. The experiments showcased the device being able to bypass the thrombus analog in all cases. The results were especially promising in case of platelet-rich thrombus analogs: no fragmentation occurred and the device was also able to remove the thrombi. This is an important observation since currently used devices are less effective when used on platelet-rich, fibrous thrombi as compared to RBC-rich thrombi [7] since platelet-rich thrombi are firmer and thus more resistant to penetration than the RBC-rich ones. This study suggests that approaching this type of thrombus along its circumference is a viable strategy. In case of the RBC-rich thrombus analogs the experiment has showcased fragmentation in one of the attempts, although relatively small volume-wise. The fragmentation might have been caused by the wire of the arms cutting through the thrombus analog: if the friction between the expanding set and the thrombus has caused the thrombus to move forward while the gripping set has remained static it could result in the grasping set shearing the thrombus. Perhaps in the next iteration the gripping element could have a shape of a filled-out semi-circle rather than its circumference to avoid cutting through the thrombus analog. In all of the experiments it was observed that the thrombus was displaced in the direction of the device’s advancement. While this did not prevent the device from bypassing the thrombus analogs it is preferable to minimize the forward push since it can prolong the duration of the intervention. The forward push is likely the result of friction between the thrombus and the arms advancing along it. In future iterations the forward push can be mitigated by increasing the grip strength which would counteract the push force. The grip strength can be increased by altering the shape and the surface texture of the grippers. Additionally the arms could be redesigned in such a way that they expand radially along their entire length, not just their ends, thus minimizing surface contact with the thrombus during the advancement phase.

While the results are promising, the experiment was subject to several limitations. The material property provided by the manufacturer of the silicone used for the phantom vessels was shore hardness. Literature on vein biomechanics provides the elastic modulus of the veins [11]. The conversion ratio between shore hardness and elastic modulus is only a rough approximation, thus the choice of material was mostly based on consultations with experienced clinicians and their tactile assessment of the phantom vessels. The phantom vessels used in the experiment were cylindrical while real vessels either taper or get wider, depending on the catheterization site relative to the site of the occlusion. Another geometric factor that was omitted in the phantom vessels is the presence of valves. We decided not to model valves because studies have shown that devices such as the Clottriever, which slide along vessel walls during removal, do not lead to valvular injury [12]. While not prone to injury, the presence of valves could affect the resistance to insertion and removal of device’s arms. While the stiffness of the phantom vessels is approximately comparable to the stiffness of real vessels, the thrombus-vessel surface adhesion was not accounted for. Additionally the thrombus analogs were inserted into the phantom vessel rather than maturing inside it, which can also affect the surface interaction between the two, especially in the case of fibrous thrombi, which can bond to vessel walls. Another limitation is the fact that the setup was not dynamic: the experiment was carried out at atmospheric pressure in the absence of any fluid flow: in biological conditions the force of blood flow and the pressure difference between the two sides of the thrombus can be one of the main forces that need to be counteracted during thrombus removal.

The prototype presented in this study is a proof of concept and adjustments have to be incorporated into the design before clinical application would come into view. In order to prevent blood vessel wall damage, the arms of the device should be made at least slightly flexible: blood vessels are generally curved and insertion of a straight, rigid object could cause injury. While the device presented in this study has been designed with DVT in mind, the principles of thrombectomy are similar in cases such as acute ischemic stroke or pulmonary embolism. Adapting the device for these applications would require adjusting the diameter of the device, enabling actuation of the device over a large distance (catheters used for stroke thrombectomy can reach up to 160 cm working length [13]) and making the device more flexible as brain vasculature is more fragile and tortuous. While these adjustments would need to be made, the working principle of the mechanism, i.e. bypassing the thrombus in small increments while expanding the vessel lumen, could be just as effective in those cases. Lastly, while the device proposed in this study is meant for bypassing the thrombus and not for removing it, our experiments have shown that in the case of platelet-rich thrombi analogs the device is capable of removal. Future work may also focus on utilizing this bypassing method in RBC-rich thrombus removal, either by integrating this device with currently used thrombectomy devices for use in pair with an aspiration catheter, or by developing the device further so that it can provide grip firm enough for removal.

## Conclusion

In this study a prototype of a novel device aiding thrombectomy has been developed and evaluated. The design utilizes an innovative strategy of combining vessel lumen expansion thus reducing normal force acting on the thrombus with incremental advancement of grippers. The prototype has been evaluated in a custom-made experimental setup using various phantom blood vessels and thrombi analogs. The experiments have shown that the device can successfully reach behind the thrombus. The prototype was more effective in the removal of the platelet-rich, fibrous thrombi, which is more challenging than RBC-rich thrombi removal with currently used devices. The proposed prototype showcases the potential of the expand-grasp-advance approach and can be a foundation for further research of this principle and next iterations of such device.

## Data Availability

All relevant data are within the manuscript and its Supporting Information files.

## Supporting information

**S1 Video. Prototype during being actuated.**

## Acknowledgments

We would like to thank Pieter Jan van Doormaal and the interventional radiology team at Erasmus MC for offering their clinical perspective on the subject as well as Sanne van Kuijk and Luca Bontempi for their help with the experiments.

